# How pain neuroscience education is performed in patients with chronic musculoskeletal pain: a scoping review protocol

**DOI:** 10.1101/2021.03.04.21252912

**Authors:** Roberta Giovinazzi, Andrea Tamborrino, Michele Marelli, Leonardo Pellicciari, Tiziano Innocenti

## Abstract

**Background:** Pain Neuroscience Education (PNE) is an effective widely used strategy in the field of chronic pain management. The objective of this scoping review is to summarize the literature to identify how PNE is performed in patients with chronic musculoskeletal pain.

**Methods:** This scoping review will be performed in accordance with the Joanna Briggs Institute methodology. Studies will be included if they meet the following population, concept, and context criteria: any patient with chronic musculoskeletal pain aged over 18 years old, any PNE delivery method, any context. No study design, publication type, and data restrictions will be applied. MEDLINE, Cochrane Central, Scopus, PsycInfo and PEDro databases will be searched up to March 2021. Additional records will be identified through searching in grey literature and reference lists of all relevant and included studies. Two reviewers will independently screen all title, abstracts and full-text studies for inclusion. A data collection form will be developed by the research team to extract the characteristics of the studies included. A tabular and accompanying narrative summary of the information will be provided.

**Conclusions:** This will be the first scoping review to provide a comprehensive overview of the topic. The results will add meaningful information for future research and clinical practice. Furthermore, any knowledge gaps on the topic will be identified. The results of this research will be published in a peer-reviewed journal and will be presented at relevant (inter)national scientific events.

## Introduction

Pain neuroscience education (PNE) is an educational approach used in the management of chronic pain.^1^ PNE aims to reconceptualise the patients’ understanding of their own pain as less threatening and reduce catastrophizing thinking to facilitate a more active coping strategy.^2^ It is based on a biopsychosocial model according to the nature of chronic pain.^1,3,4^

PNE has been widely implemented and tested in the field of chronic musculoskeletal pain (CMP). Recent reviews, investigating the effectiveness of PNE for adults with CMP, found that PNE produces clinically important reductions on pain catastrophizing and kinesiophobia in the short and medium term,^2,5^ consequently on pain and disability.^2,6,7^ Despite evidence supports PNE effectiveness, it is only one component of an overall management strategy for chronic pain.^8^ In fact, education as a stand-alone treatment has minimal effects on pain and disability.^2,5^ A significant effect can be observed in patients with CMP when PNE is delivered in combination with other interventions, such as exercise and physical activity.^2^ Another important factor in determining PNE effectiveness is its relevance to each individual, for instance tailoring the educational material on individuals’ needs.^9,10^

Moreover, additional effects have been found on the longer duration of PNE, but the precise dosage is unknown.^5^ In 2011, Jo Nijs provided practical guidelines about how to perform PNE,^1^ but several features of its administration, such as dosage, format, structure, still need to be standardized. Despite some aspects such as contextual factors cannot be standardized, investigating around this matter is necessary as authors of PNE systematic reviews underline great heterogeneity among the included studies.^2,5–7^ They refer to the studies’ methodology, the type of participants, interventions, comparisons, outcomes, and the different modalities by which PNE is performed. This heterogeneity could be considered a limitation both for research and clinical practice.

Therefore, the objective of this project is to summarize the literature to identify how PNE is performed in patients with CMP. This objective corresponds to the purpose of a scoping review study design. As maintained by the Joanna Briggs Institute (JBI),^11^ a scoping review approach may be used to map and clarify key concepts, identify gaps in the research knowledge base, and report on the types of evidence that address and inform practice in the field. Scoping reviews are particularly helpful when the literature is complex and heterogeneous.^12^

In particular, the objectives of this study will be to:

1. Provide a comprehensive overview of all studies dealing with PNE delivery methods.
2. Identify and summarize studies according to the type of delivery method, with specific subgroups that will emerge from the analysis of the included studies.
3. Identify any knowledge gaps on the topic.
4. Share review findings with the scientific community.

## Methods

The proposed scoping review will be conducted in accordance with the Joanna Briggs Institute methodology (JBI) for scoping reviews.^11,12^ The Preferred Reporting Items for Systematic reviews and Meta-Analyses extension for Scoping Reviews (PRISMA-ScR) Checklist for reporting will be used.^13^

### Research question

We formulated the following research question: “How is PNE performed in patients with chronic musculoskeletal pain in the existing literature?”

### Inclusion criteria

#### Population

This review will consider eligible studies that (1) include any chronic musculoskeletal pain conditions, (2) include pain that existed for 3 months or more, (3) only examined an adult population (18 years and older).

#### Concept

This review will consider eligible studies that include PNE as part of the intervention.

#### Context

This review will include studies conducted in any context.

#### Sources

This scoping review will consider any study designs or publication type for inclusion. No time, geographical, and setting restrictions will be applied. Studies published in English or Italian language will be included.

Studies that do not meet the above-stated Population-Concept-Context (PCC) criteria or that provide insufficient information will be excluded.

### Search strategy

An initial limited search of MEDLINE was undertaken to identify articles on the topic. The words contained in the titles and abstracts of the relevant articles and the index terms used to describe PNE were used to develop a full search strategy for MEDLINE (see ***Table 1***, which shows the search strategy for each database). The search strategy, including all identified keywords and index terms, will be adapted for use in Cochrane Central, Scopus, PsycInfo and PEDro. In addition, also grey literature (e.g., Google scholar, direct contact with experts in the field of PNE and chronic pain) and the reference lists of all relevant and included studies will be searched. The complete search strategy will be reported according to the PRISMA-S^14^ (extension for reporting literature searches in systematic reviews) and it will be detailed in a supplementary file.

**Table 1.** Search strategy for each database.

| Database | Search strategy |
| --- | --- |
| MEDLINE | ("pain neuroscience education" OR "therapeutic neuroscience education" OR "pain biology education" OR "pain physiology education" OR "pain neurophysiology education" OR "explaining pain" OR "explain pain" OR "pain education" OR "pain science education" OR "neuroscience education" OR pain education[Title/Abstract]) |
| Cochrane Library | ("pain neuroscience education" OR "therapeutic neuroscience education" OR "pain biology education" OR "pain physiology education" OR "pain neurophysiology education" OR "explaining pain" OR "explain pain" OR "pain education" OR "pain science education" OR "neuroscience education" OR "pain education") in All Text |
| PsychINFO | ("pain neuroscience education" OR "therapeutic neuroscience education" OR "pain biology education" OR "pain physiology education" OR "pain neurophysiology education" OR "explaining pain" OR "explain pain" OR "pain education" OR "pain science education" OR "neuroscience education" OR "pain education") |
| Scopus | TITLE-ABS-KEY ( "pain neuroscience education" OR "therapeutic neuroscience education" OR "pain biology education" OR "pain physiology education" OR "pain neurophysiology education" OR "explaining pain" OR "explain pain" OR "pain education" OR "pain science education" OR "neuroscience education" OR "pain education" ) |
| PEDro | "pain education"<br>"neuroscience education"<br>"biology education"<br>"physiology education"<br>"neurophysiology education"<br>"explaining pain"<br>"explain pain" |

### Study selection

Once the search strategy has been successfully completed, search results will be collected and imported into EndNote X7 (Clarivate Analytics, PA, USA). Duplicates will be automatically removed before the file containing a set of unique records is made available to reviewers for further processing (i.e., study screening and selection). The selection process will consist of two levels of screening using Rayyan QCRI online software:^15^ (1) a title and abstract and (2) a full-text screening. For both levels, two investigators (RG and MM) will screen the articles independently to determine if they meet the inclusion/exclusion criteria. If there are disagreements, they will be resolved through discussion or a third reviewer (LP). Reasons for the exclusion of each record will be recorded and reported in the supplementary materials. The entire selection process will be reported and presented in a PRISMA flow diagram.^16^

### Data extraction

An ad-hoc data extraction form will be developed by the reviewers and key information on the selected articles, such as population, concept and context, will be collected. A draft extraction tool is provided (see ***Table 2***, which illustrates the PNE delivery methods data extraction instrument). This form will be reviewed by the research team and pre-tested by all reviewers before implementation to ensure that the form captures the information accurately. Charting results is commonly an iterative process during scoping reviews; other data can be added to this form according to the subgroups that could emerge from the analysis of the studies included. Modifications will be detailed in the full scoping review.

**Table 2.** PNE delivery methods data extraction instrument.

| Author,<br>year | Educational delivery methods |  |  |  |  |  |  |  |  |  |
| --- | --- | --- | --- | --- | --- | --- | --- | --- | --- | --- |
|  | Associated<br>intervention | Individual | Number<br>of<br>sessions | Duration/<br>session | Format | Content | Use of teaching tools<br>(examples, metaphors,<br>images, video, ppt) | Use of<br>specific test | Written<br>information to<br>read at home | Provider<br>(clinician<br>training) |

### Data management

As a scoping review, the purpose of this study is to aggregate the findings and present an overview on the research rather than to evaluate the quality of the single studies. The results will be presented in two ways:

1. Numerically. Data extraction will be summarized in tabular form. Further categories may be added if considered appropriate.
2. Thematically. A descriptive analysis will be performed pertaining to themes and key concepts relevant to the research questions and according to subgroups that could emerge.

## Ethics and dissemination

A manuscript with results will be prepared and submitted for journal publication upon project completion. The findings of the study will be disseminated at a relevant (inter)national conference. The results of this research will be published in a relevant peer-reviewed journal in the rehabilitation and physical therapy field. All results of this scoping review will also be announced at (inter)national scientific events in the area of rehabilitation of musculoskeletal disorders.

## Data Availability

Not applicable

## Acknowledgements

Not applicable.

## List of abbreviations

PNE: Pain Neuroscience Education
CMP: Chronic Musculoskeletal Pain
PCC: Population-Concept-Context
JBI: Joanna Briggs Institute
PRISMA: Preferred Reporting Items for Systematic Reviews and Meta-Analyses

## List of additional files

***Table 1***: shows the search strategy for each database.

***Table 2***: illustrates the PNE delivery methods data extraction instrument.

## References

1. Nijs, J., Van Wilgen, C. P., Van Oosterwijck, J., van Ittersum, M. & Meeus, M. How to explain central sensitization to patients with ‘unexplained’ chronic musculoskeletal pain: practice guidelines. Man. Ther. 16, 413–418 (2011).

2. Louw, A., Zimney, K., Puentedura, E. J. & Diener, I. The efficacy of pain neuroscience education on musculoskeletal pain: a systematic review of the literature. Physiother. Theory Pract. 32, 332–355 (2016).

3. Moseley, G. L. & Butler, D. S. Fifteen years of explaining pain: the past, present, and future. J. Pain 16, 807–813 (2015).

4. Linton, S. J. & Shaw, W. S. Impact of psychological factors in the experience of pain. Phys. Ther. 91, 700–711 (2011).

5. Watson, J. A. et al.. Pain neuroscience education for adults with chronic musculoskeletal pain: a mixed-methods systematic review and meta-analysis. J. Pain (2019).

6. Louw, A., Diener, I., Butler, D. S. & Puentedura, E. J. The effect of neuroscience education on pain, disability, anxiety, and stress in chronic musculoskeletal pain. Arch. Phys. Med. Rehabil. 92, 2041–2056 (2011).

7. Wood, L. & Hendrick, P. A. A systematic review and metalJanalysis of pain neuroscience education for chronic low back pain: ShortlJand longlJterm outcomes of pain and disability. Eur. J. Pain 23, 234–249 (2019).

8. Society, B. P. Guidelines for pain management programmes for adults. An evidence-based Rev. Prep. behalf Br. Pain Soc. (2013).

9. Robinson, V., King, R., Ryan, C. G. & Martin, D. J. A qualitative exploration of people’s experiences of pain neurophysiological education for chronic pain: The importance of relevance for the individual. Man. Ther. 22, 56–61 (2016).

10. King, R. et al. Pain reconceptualisation after pain neurophysiology education in adults with chronic low back pain: A qualitative study. Pain Res. Manag. 2018, (2018).

11. Peters, M. et al. The Joanna Briggs Institute reviewers’ manual 2015: methodology for JBI scoping reviews. (2015).

12. Peters, M. D. J. et al. Updated methodological guidance for the conduct of scoping reviews. JBI Evid. Synth. 18, 2119–2126 (2020).

13. Tricco, A. C. et al. PRISMA extension for scoping reviews (PRISMA-ScR): checklist and explanation. Ann. Intern. Med. 169, 467–473 (2018).

14. Rethlefsen, M. L. et al. PRISMA-S: an extension to the PRISMA Statement for Reporting Literature Searches in Systematic Reviews. Syst. Rev. 10, 1–19 (2021).

15. Ouzzani, M., Hammady, H., Fedorowicz, Z. & Elmagarmid, A. Rayyan—a web and mobile app for systematic reviews. Syst. Rev. 5, 1–10 (2016).

16. Moher, D., Liberati, A., Tetzlaff, J., Altman, D. G. & Group, P. Preferred reporting items for systematic reviews and meta-analyses: the PRISMA statement. PLoS med 6, e1000097 (2009).

